# Occupational stigma and post-traumatic stress disorder among healthcare workers

**DOI:** 10.64898/2025.12.15.25342262

**Authors:** Charline Vincent, Roberto Mediavilla, Honor Scarlett, Wissam El-Hage, Pierre Chauvin, Cécile Vuillermoz

## Abstract

**Background:** Healthcare workers (HCWs) could be subject to stigma, particularly in contexts of fear of contagion related to various communicable diseases. While existing literature has established links between workplace stigma and adverse mental health outcomes, this research has largely focused on stigma derived from personal characteristics (e.g., race, gender). However, studies specifically investigating occupational stigma - the stigma resulting directly from the nature of the healthcare profession itself - remain scarce.

**Objective:** We investigated the association between perceived occupational stigma and post-traumatic stress disorder (PTSD) symptoms one year following the onset of the COVID-19 pandemic.

**Methods:** This study analyzed data from a cross-sectional online survey conducted in France. PTSD symptoms were measured with the Posttraumatic Stress Disorder Checklist (PCL-5), and perceived stigma was assessed with a single-item question. Associations were analyzed using inverse probability weighting based on propensity scores, which accounted for a broad range of potential confounders, including socioeconomic factors, work characteristics, and comorbidities.

**Results:** Among 655 respondents, who were mainly physicians, 44.8% reported experiencing occupational stigma and 8.7% met the threshold for PTSD symptoms. After adjustment, perceived occupational stigma was significantly associated with PTSD symptoms (ORa=2.16; 95% CI [1.09–4.25]).

**Conclusion:** Occupational stigma is independently associated with PTSD symptoms among HCWs. These findings underscore the critical need for integrating targeted anti-stigma interventions into mental health strategies for HCWs involving in managing infectious disease or in pandemic context.

## Introduction

During the COVID-19 pandemic, in addition to the increased pressure on the healthcare system to manage the health crisis, healthcare professionals faced various reactions from the population. One of these was the expression of gratitude toward healthcare workers (HCWs), or even their heroization at the height of the crisis (Bauchner et al., 2020). However, opposite reaction were also expressed, such as fear and rejection (Faghani et al., 2023). Some studies reported that HCWs were perceived as a source of infection, and therefore were sometimes the target of insults, verbal abuse, or rejection, due to fear of contamination (D’Alessandro et al., 2022). In addition, some studies also showed that HCWs were targeted by people dissatisfied with the healthcare system’s handling of the COVID-19 crisis, or refusing the protective measures made mandatory by public authorities, and, later, vaccination recommendations (Sari et al., 2023).

These attitudes can be perceived as acts of stigmatization by HCWs. Stigmatization is a powerful social process characterized by labeling, stereotyping, and rejection of an individual, leading to loss of status and discrimination, all within a context of power imbalance (Link & Phelan, 2001). More specifically, it refers to social/public stigmatization when it is based on a belief shared by a large part of society and is embedded in the social framework to create a sense of inferiority (Ahmedani, 2011). Social stigmatization of HCWs has been documented following successive epidemics (Wester & Giesecke, 2019) and finally during the COVID-19 pandemic, where HCWs reported feeling stigmatized because of their profession (Bagcchi, 2020). Stigma can be very disempowering, as it often leads to rejection from social circles communities (Lin et al., 2007) and reduces self-esteem and a sense of belonging (Chamaa et al., 2021). It can increase the risk of developing or worsening mental health disorders, such as depression and anxiety disorders (Lewis et al., 2022). Some studies have indicated that the perceived experience of occupation-related stigma was also a risk factor for posttraumatic stress disorder (PTSD) in HCWs (Bryant-Genevier et al., 2021; Narita et al., 2022).

Numerous studies have observed a high level of mental health disorders among HCWs during the COVID-19 pandemic (Y. Li et al., 2021). Some meta-analyses have highlighted high levels of PTSD symptoms among HCWs in 2020, ranging from 21% (Y. Li et al., 2021) to 31% (Marvaldi et al., 2021). One year after the onset of the pandemic, the prevalence of PTSD ranged from 13% to 37% in cross-sectional observational studies (Bahadirli & Sagaltici, 2021; Bryant-Genevier et al., 2021; Carola et al., 2022; Esteban-Sepúlveda et al., 2022). PTSD symptoms among HCWs can have significant consequences. Individuals experiencing PTSD symptoms are at higher risk of developing other mental health conditions, such as depression and anxiety (Rossi et al., 2021), which may have a direct impact on both their personal and professional lives. In professional life, mental disorders can increase absenteeism but also presenteeism (being at work despite poor health), which can lead HCWs to more medical errors and poorer quality of care (Braquehais & Vargas-Cáceres, 2023).

To examine the relationship between occupational stigma and PTSD, potential confounders should be considered. Indeed, the literature suggests that many social, living, working and health determinants are related to one or the other (or for some, both). Several social determinants of PTSD among HCWs have been identified during the COVID-19 pandemic, such as being a woman (Costa et al., 2023), young age (Marsden et al., 2022; Rossi et al., 2021), having children (Jiang et al., 2022), low level of education (D’Alessandro-Lowe et al., 2024), low income (Bryant-Genevier et al., 2021; Costa et al., 2023), financial difficulties (Boitet et al., 2023), and lack of social support at work (Marsden et al., 2022) or from family and friends (Cénat et al., 2022; Costa et al., 2023). It has also been shown that working conditions during the COVID-19 pandemic may have increased the risk of experiencing PTSD symptoms depending on factors such as the type of profession (nurse, doctor, or other) (Bahadirli & Sagaltici, 2021; Costa et al., 2023), seniority in the position held (Bahadirli & Sagaltici, 2021), the number of changes in work schedules (Bryant-Genevier et al., 2021) and frontline duties (Rossi et al., 2021), night work (Cousin Cabrolier et al., 2023) or work in areas with overcrowded beds (Bonzini et al., 2022), being forced to change positions (Rigas et al., 2024), the type of healthcare structure (Ouazzani Housni Touhami et al., 2023), and being a student or trainee (Wilgenbusch et al., 2023). From a work organization perspective, lack of awareness of psychosocial risks or stress management training is also associated with PTSD symptoms (Motreff et al., 2020). Additionally, the literature has demonstrated a relationship between PTSD symptoms and the perceived level of exposure to the pandemic (Bryant-Genevier et al., 2021), having been previously diagnosed with COVID-19 (Bahadirli & Sagaltici, 2021), and high COVID-19 vaccination rates (Hsieh et al., 2022). Lastly, some health comorbidities are known to be associated with PTSD symptoms, such as depression and anxiety (Rossi et al., 2021), burnout (P. Li et al., 2021), poor resilience (Costa et al., 2023), history of trauma (Tebbeb et al., 2022), history of psychiatric medication (Alshehri et al., 2023), having received recent psychological support (Rossi et al., 2021), and chronic somatic diseases (Bahadirli & Sagaltici, 2021).

On the other hand, not only can stigmatizing processes affect multiple aspects of individuals’ lives, but also, as social processes, they are likely to target the most socially vulnerable individuals (Link & Phelan, 2001). Indeed, many social characteristics can be associated with stigma, such as lack of social support (Minichil et al., 2021), being a woman (Minichil et al., 2021) and holding a low-skilled job (Mostafa et al., 2020).

For the first and only time in France, the objective of this study was to examine, in a voluntary sample of HCWs, whether occupational stigma was associated with PTSD symptoms one year after the start of the COVID-19 pandemic, taking into account socioeconomic status, social support, working conditions, exposure to COVID-19 and health comorbidities, and using an original analysis approach based on machine learning methods to generate propensity scores.

## Methods

### Study design and data collection

This study is based on data collected by the French PSYCOVER survey (PSYchological impact of the COVID-19 pandemic in healthcare workERs) (Bertuzzi et al., 2022). This online survey on volunteers was conducted between April and October 2021, i.e. 13-months after the first surge of COVID-19 cases in France (Costemalle et al., 2021) and coinciding with the fourth peak of the pandemic. During May and June 2021, a curfew was enforced, and numerous establishments open to the public remained closed. Subsequently, in July 2021, a “health pass”, requiring proof of a negative PCR test within the last 72 hours or full vaccination, was put in place to allow access to many public venues.

Inclusion criteria in PSYCOVER survey were being an adult who had work in a healthcare facility that had received patients with COVID-19 since the beginning of the pandemic, speaking one of the study languages (English and French), and having given an informed consent to participate. Students and trainees were also eligible to participate. All types of HCWs could participate, i.e. – according to the French Public Health Code (LegiFrance, 2023) - medical professionals (physicians, midwives, dentists), pharmacists, medical physicists, and medical assistants (nurses, assistant nurses, childcare assistants, paramedics, physiotherapists, occupational therapists etc.).

Participants were recruited through: (1) calls to participate in social media platforms such as Facebook, Twitter, and LinkedIn; (2) communication with scholarly societies and professional associations (200 were contacted), asking them to inform and refer their members to the study website; (3) emails to institutional and academic partners; and (4) information to the managers of the 32 French university hospitals. Our final study population consisted of 655 HCWs.

The study protocol received approval from the Sorbonne University Ethical Committee (No. 2020-CER-2020-27) and was reported to the French Commission on Information Technology and Liberties, CNIL (No. 2222413, 20-05-2021).

### Measures

#### Occupational stigma

Participants were asked the following question with a yes, unsure, or no response option: “*Since the start of the COVID-19 pandemic, have you felt that you have been treated differently and unfairly because of your occupation as a healthcare professional by friends or family you live with, other friends, other family members, neighbors, parents of children in your community, or other people?*”. Those who answered “yes” or “unsure” were considered to “have perceived stigma related to their profession” and those who answered “no” as “having not perceived stigma related to their profession”.

#### Outcome

PTSD symptoms were assessed with the Posttraumatic Stress Disorder Checklist (PCL-5), a 20-item self-reported measure corresponding to DSM-5 symptom criteria for PTSD (Weathers et al., 2013), validated in French (Ashbaugh et al., 2016). The PCL-5 includes four subscales describing DSM-5 clusters corresponding to the re-experiencing negative events, avoidance, negative cognition and mood, and arousal. Responses to each item were collected with Likert scales ranging from 0 (not at all) to 4 (extremely), with a total score ranging from 0 to 80. We tested the distribution of the variable using Shapiro-Wilk tests. As it did not follow a normal distribution, we dichotomized it. We considered that scores ≥33 indicated a positive screening for PTSD (Weathers et al., 2013).

#### Covariates

Based on the existing scientific literature, we considered covariates associated with PTSD symptoms and/or stigma.

#### Sociodemographic characteristics

Gender, age (under 40 years old vs. over 40, considering the median age of the study population), self-reported monthly household income (dichotomized according to its distribution as >4000 € (first quartile) or ≤4000 €), educational level (below MD or PhD, versus higher), having children under 3 years old (yes/no) and perceived financial evolution since the beginning of the pandemic (deteriorated/not changed or improved) were taken into account.

#### Social support

Social support was assessed in 4 dimensions, each with a specific question: 1) “*Do you know any key person who could support you within your institution?*” (yes/no) for psychosocial support at work; then “*Since the beginning of the pandemic, if needed, could you count on anyone, whether they were members of your household, other family members, friends or neighbors, colleagues, community members to*…” 2) *“Provide you with moral or emotional support?”* (yes/no) for moral support; 3) *“Help you financially or materially?”* (yes/no) for material and financial support; 4) *“Help you in your daily life, give you a hand?”* (yes/no) for support in daily life. We also asked whether the respondent lived alone or not.

#### Working conditions

We examined whether the following characteristics were associated with PTSD symptoms: occupation (nurse, physician, other); length of time in current job (less than 6 months vs. more); a change in work hours since the beginning of the pandemic (increased vs. no change or decreased); having been forced to change position (role, department or specialty) since the beginning of the pandemic; having worked in a geographical area in which the epidemic was severe (yes/no), i.e. if overcrowding of hospital beds was observed during at least two epidemic waves; working night shifts (often or always/sometimes or never); place of practice (hospital versus ambulatory practice); working in intensive care or an emergency unit (yes/no); type of employment contract (temporary versus fixed-term contract or tenure position); being a student or a trainee (yes/no); teamwork (yes/no), and having a supervisor at work (yes/no).Psychosocial risk awareness at work was measured using the question: *“Have you been informed about psychosocial risks in the context of your professional activities (excessive stress, psychological trauma, burnout, etc.) through specific training?”* (yes/no). Stress management training was measured using the question: *“Have you been trained to manage your stress in such situations?”* (yes/no).

#### Exposure to COVID-19

Exposure to COVID-19 was assessed through the perceived exposure to COVID-19 (rated on a scale from 0 to 10, with 10 indicating very high exposure, and using a median cut-off of 7); having been tested positive for COVID-19 (at least once at the time of the survey); having had relatives hospitalized for COVID-19 and having been a full vaccination for COVID-19.

#### Health comorbidities

Depressive symptoms were assessed using the 9-item Patient Health Questionnaire (PHQ-9) (Kroenke & Spitzer, 2002) (a score ≥10 was considered to be indicative of the presence of depressive symptoms (Kroenke et al., 2001)). Anxiety symptoms were assessed using the 7-item Generalized Anxiety Disorder Scale (GAD-7) (a score ≥10 was considered to be indicative of the presence of anxiety symptoms (Spitzer et al., 2006)). Burnout was assessed using the Maslach Burnout Inventory test (MBI) (Maslach et al., 1996) which has been validated in French (Faye Dumanget et al., 2015). We chose to dichotomize burnout into two categories (yes/no) based on the method proposed by Maslach which considers burnout to be characterized by high emotional exhaustion (score>29) and low personal accomplishment (score ≤33), or high emotional exhaustion (score>29) and high depersonalization (score >11) (Maslach et al., 1996). Resilience was assessed using Connor-Davidson Resilience Scale-10 (CD-RISC-10) (Connor & Davidson, 2003) (using the first quartile, we considered a score <22 indicative of the absence of resilience). Past experience of a traumatic event within the past 12 months (yes/no), history of psychotropic treatment for more than 6 months before the pandemic (yes/no), history of major lifetime somatic health problems (yes/no) and psychological support related to the pandemic (yes/no) were also measured.

## Statistical analysis

Statistical analyses were carried out by using R Studio version 4.3.2. The PsyCOVer survey used a non-probabilistic sampling method. Approximately 70% of survey respondents were physicians - although they represent only 18% of HCWs in France - and 42% worked in ambulatory practice – compared to 26% among French HCWs (INSEE, 2020). To reduce selection bias, we implemented a data calibration on occupation (physicians vs. other HCWs) and sector of activity (ambulatory care practice vs. hospital). We applied the margin adjustment method (Olivier Sautory, 2018), using data from the second and third quarters of the 2021 French Employment Survey (INSEE, 2023) conducted by INSEE, using the Icarus package (Rebecq, 2016).

To handle missing data (up to 8% per variable, for a total of 2.6% missing data overall), we imputed 20 data sets using the *mice* and *survey* R packages, which also allows for calibration weights to be taken into account. Linearity of discrete variables was assessed using Shapiro-Wilk tests.

Because HCWs who have been stigmatized due to their profession are likely to differ from those who have not on a number of characteristics (Claustre et al., 2020), we calculated propensity scores for occupational stigma. All analyses were conducted on imputed and weighted datasets. We first ran univariate logistic regression models to test associations between covariates and occupational stigma and between covariates and PTSD symptoms. All variables, whether or not associated with the study outcome, were included. If covariates were only associated with the study exposure, they were excluded from further analyses: indeed, including variables associated with the exposure but not the outcome increases the variance of the estimated effect without reducing bias (Brookhart et al., 2006). Second, we used generalized boosted models (GBM), a powerful form of machine learning to generate propensity scores (Harder et al., 2010). We chose the inverse probability weighting (IPW) method implemented by *weightThem* function (Pishgar et al., 2021), which, unlike other techniques, has the advantage of preserving the entire initial sample for analysis (Dong et al., 2020). Third, we checked the weights validity indicators. To estimate the adequate balance of covariate across exposure groups, we used the Absolute Standard Mean Difference (ASMD) under 0.2 (McCaffrey et al., 2013) and Kolmogorov-Smirnov (KS) under 0.1 (McCaffrey et al., 2013). A visual plot display (Noah Greifer, 2023) enabled us to exclude variables that did not have good adjustment criteria. Fourth, we multiplied the propensity scores obtained by the previously calculated survey weights to fit an IPW-logistic regression model and estimate the association between stigma and PTSD effect of study exposure on the outcome of interest, expressed as an aOR and its 95% CI. A p value <0.05 indicated statistical significance.

We also performed a sensitivity analysis using the same approach but where the experience of perceived occupational stigma was determined only from a “yes” response to the corresponding question while participants who answered “unsure” were excluded from the analysis.

## Results

Descriptive statistics of the total sample were first conducted on the raw dataset (Table 1). Among our sample of HCWs, 74.8% were female, and 48.0% were under 40 years old. Two thirds (65.6%) had a level of education higher than or equal to MD or PhD. The sample comprised 71.6% physicians, 11.1% nurses, and 17.3% from other occupations. The majority (57.9%) worked in hospitals, and 17.4% in intensive care or emergency units. We observed that 44.8% of participants reported experiencing a occupational stigma during the COVID-19 pandemic and that 53 participants (8.7%) exhibited symptoms of PTSD.

**Table 1.** Description of the study population

|  |  | Raw data |  |  |  |  |  | Weighted data |  |  |  |  |  |
| --- | --- | --- | --- | --- | --- | --- | --- | --- | --- | --- | --- | --- | --- |
|  |  | Total |  | Occupational stigma |  | PTSD symptoms |  | Total |  | Occupational stigma |  | PTSD symptoms |  |
|  |  | n = 655 |  | n = 278 |  | n = 53 |  | n = 1418 |  | n = 625 |  | n = 129 |  |
|  |  | n | % | n | % | n | % | n | % | n | % | n | % |
| **Experience of occupational stigma** | No stigmatization | 343 | 55.2 |  |  | 15 | 29.4 | 724 | 53.7 |  |  | 34 | 26.8 |
|  | Probable stigmatization | 278 | 44.8 |  |  | 36 | 70.6 | 625 | 46.3 |  |  | 93 | 73.2 |
| **Socio-demographic** |  |  |  |  |  |  |  |  |  |  |  |  |  |
| Gender | Male | 164 | 25.2 | 60 | 21.8 | 9 | 17.0 | 306 | 21.7 | 113 | 18.2 | 19 | 14.7 |
|  | Female | 487 | 74.8 | 215 | 78.2 | 44 | 83.0 | 1105 | 78.3 | 507 | 81.8 | 110 | 85.3 |
| Age | Over 40 years old | 335 | 52.0 | 142 | 52 | 24 | 45.3 | 728 | 52.3 | 329 | 53.8 | 60 | 46.5 |
|  | Under 40 years old | 309 | 48.0 | 131 | 48 | 29 | 54.7 | 663 | 47.7 | 283 | 46.2 | 69 | 53.5 |
| Monthly household income | > 4000 euros | 405 | 64.8 | 162 | 61.4 | 24 | 47.1 | 794 | 58.9 | 323 | 54.7 | 55 | 43.7 |
|  | ≤ 4000 euros | 220 | 35.2 | 102 | 38.6 | 27 | 52.9 | 554 | 41.1 | 267 | 45.3 | 71 | 56.3 |
| Educational level | ≥ MD or PhD | 430 | 65.6 | 175 | 62.9 | 25 | 47.2 | 718 | 50.6 | 293 | 46.9 | 43 | 33.3 |
|  | < MD or PhD | 225 | 34.4 | 103 | 37.1 | 28 | 52.8 | 700 | 49.4 | 332 | 53.1 | 86 | 66.7 |
| Having children | No | 537 | 83.1 | 97 | 35.0 | 28 | 52.8 | 529 | 37.7 | 222 | 35.6 | 61 | 47.3 |
|  | Yes | 109 | 16.9 | 180 | 65.0 | 25 | 47.2 | 875 | 62.3 | 401 | 64.4 | 68 | 52.7 |
| Perceived financial evolution SBP<sup>1</sup> | Not changed or improved | 566 | 86.8 | 229 | 82.4 | 42 | 79.2 | 1225 | 86.7 | 522 | 83.5 | 105 | 81.4 |
|  | Deteriorated | 86 | 13.2 | 49 | 17.6 | 11 | 20.8 | 188 | 13.3 | 103 | 16.5 | 24 | 18.6 |
| **Social support** |  |  |  |  |  |  |  |  |  |  |  |  |  |
| Perceived psychosocial support at work | Yes | 340 | 52.6 | 149 | 53.8 | 20 | 37.7 | 770 | 54.9 | 345 | 55.6 | 59 | 45.7 |
|  | No | 307 | 47.4 | 128 | 46.2 | 33 | 62.3 | 633 | 45.1 | 276 | 44.4 | 70 | 54.3 |
| Perceived moral support | Yes | 592 | 92.8 | 248 | 90.2 | 41 | 77.4 | 1279 | 92.5 | 556 | 89.7 | 101 | 78.3 |
|  | No | 46 | 7.2 | 27 | 9.8 | 12 | 22.6 | 104 | 7.5 | 64 | 10.3 | 28 | 21.7 |
| Perceived financial support | Yes | 512 | 80.5 | 211 | 77.0 | 32 | 60.4 | 1070 | 77.6 | 441 | 71.6 | 77 | 59.7 |
|  | No | 124 | 19.5 | 63 | 23.0 | 21 | 39.6 | 308 | 22.4 | 175 | 28.4 | 52 | 40.3 |
| Perceived social support in daily life | Yes | 529 | 83.0 | 215 | 78.2 | 29 | 54.7 | 1124 | 81.3 | 466 | 75.2 | 71 | 55 |
|  | No | 108 | 17.0 | 60 | 21.8 | 24 | 45.3 | 258 | 18.7 | 154 | 24.8 | 58 | 45 |
| Living alone | No | 520 | 80.4 | 220 | 79.7 | 35 | 67.3 | 1109 | 79.2 | 481 | 77.7 | 87 | 68.5 |
|  |  | Total |  | Occupational stigma |  | PTSD symptoms |  | Total |  | Occupational stigma |  | PTSD symptoms |  |
|  |  | n = 655 |  | n = 278 |  | n = 53 |  | n = 1418 |  | n = 625 |  | n = 129 |  |
|  |  | n | % | n | % | n | % | n | % | n | % | n | % |
|  | Yes | 127 | 19.6 | 56 | 20.3 | 17 | 32.7 | 292 | 20.8 | 138 | 22.3 | 40 | 31.5 |
| **Working conditions** |  |  |  |  |  |  |  |  |  |  |  |  |  |
| Occupation | Others occupation | 113 | 17.3 | 42 | 15.1 | 9 | 16.9 | 335 | 23.6 | 127 | 20.3 | 28 | 21.7 |
|  | Physician | 469 | 71.6 | 193 | 69.4 | 34 | 64.2 | 798 | 56.3 | 330 | 52.8 | 62 | 48.1 |
|  | Nurse | 73 | 11.1 | 43 | 15.5 | 10 | 18.9 | 285 | 20.1 | 168 | 26.9 | 39 | 30.2 |
| Length of time in current job | More than 6 months | 558 | 86.4 | 238 | 86.5 | 40 | 76.9 | 1202 | 85.9 | 534 | 86.5 | 102 | 80.3 |
|  | Less than 6 months | 88 | 13.6 | 37 | 13.5 | 12 | 23.1 | 197 | 14.1 | 83 | 13.5 | 25 | 19.7 |
| Increased working hours SBP<sup>1</sup> | No | 221 | 33.8 | 75 | 27.0 | 14 | 26.4 | 477 | 33.7 | 159 | 25.4 | 26 | 20.2 |
|  | Yes | 432 | 66.2 | 203 | 73.0 | 39 | 73.6 | 937 | 66.3 | 466 | 74.6 | 103 | 79.8 |
| Having been forced to change position SBP<sup>1</sup> | No change | 490 | 76.0 | 197 | 71.4 | 30 | 56.5 | 1035 | 74 | 419 | 67.5 | 73 | 56.6 |
|  | At least once | 155 | 24.0 | 79 | 28.6 | 23 | 43.4 | 363 | 26 | 202 | 32.5 | 56 | 43.4 |
| Having worked in a geographical area in which the epidemic was severe | No | 208 | 33.2 | 81 | 30.5 | 16 | 32.0 | 430 | 31.8 | 174 | 29.2 | 36 | 30.3 |
|  | Yes | 419 | 66.8 | 185 | 69.5 | 34 | 68.0 | 923 | 68.2 | 421 | 70.8 | 83 | 69.7 |
| Working night shifts | Never or sometimes | 569 | 87.5 | 242 | 88.0 | 46 | 86.8 | 1216 | 86.4 | 542 | 87.6 | 115 | 89.1 |
|  | Often or always | 81 | 12.5 | 33 | 12.0 | 7 | 13.2 | 192 | 13.6 | 77 | 12.4 | 14 | 10.9 |
| Place of practice | Ambulatory practice | 274 | 42.1 | 109 | 39.4 | 15 | 28.3 | 468 | 33.3 | 185 | 29.8 | 28 | 21.7 |
|  | Hospital | 377 | 57.9 | 168 | 60.6 | 38 | 71.7 | 939 | 66.7 | 436 | 70.2 | 101 | 78.3 |
| Working in intensive care or emergency unit | No | 541 | 82.6 | 224 | 80.6 | 40 | 75.5 | 1164 | 82.1 | 503 | 80.5 | 97 | 75.2 |
|  | Yes | 114 | 17.4 | 54 | 19.4 | 13 | 24.5 | 254 | 17.9 | 122 | 19.5 | 32 | 24.8 |
| Type of employment contract | Fixed-term contract or tenure position | 296 | 47.7 | 137 | 52.1 | 26 | 52.0 | 782 | 58.1 | 374 | 63.1 | 76 | 61.8 |
|  | Temporary | 324 | 52.3 | 126 | 47.9 | 24 | 48.0 | 564 | 41.9 | 219 | 36.9 | 47 | 38.2 |
| Student or trainee | No | 575 | 88.3 | 246 | 88.8 | 46 | 86.8 | 1256 | 89 | 561 | 90 | 115 | 89.1 |
|  | Yes | 76 | 11.7 | 31 | 11.2 | 7 | 13.2 | 155 | 11 | 62 | 10 | 14 | 10.9 |
| Teamwork | Yes | 572 | 89.5 | 248 | 90.2 | 48 | 90.6 | 1264 | 91.3 | 569 | 91.8 | 117 | 90.7 |
|  | No | 67 | 10.5 | 27 | 9.8 | 5 | 9.4 | 121 | 8.7 | 51 | 8.2 | 12 | 9.3 |
| Having a supervisor at work | Yes | 339 | 53.1 | 158 | 57.5 | 35 | 66.0 | 858 | 61.9 | 407 | 65.6 | 94 | 72.9 |
|  | No | 300 | 46.9 | 117 | 42.5 | 18 | 34.0 | 527 | 38.1 | 213 | 34.3 | 35 | 27.1 |
| Psychosocial risk awareness at work | Yes | 199 | 30.4 | 76 | 27.3 | 15 | 28.3 | 421 | 29.7 | 171 | 27.4 | 39 | 30.2 |
|  | No | 455 | 69.6 | 202 | 72.7 | 38 | 71.7 | 995 | 70.3 | 454 | 72.6 | 90 | 69.8 |
|  |  | Total |  | Occupational stigma |  | PTSD symptoms |  | Total |  | Occupational stigma |  | PTSD symptoms |  |
|  |  | n = 655 |  | n = 278 |  | n = 53 |  | n = 1418 |  | n = 625 |  | n = 129 |  |
|  |  | n | % | n | % | n | % | n | % | n | % | n | % |
| Training at work to manage stress | Yes | 86 | 13.2 | 36 | 13.0 | 7 | 13.2 | 196 | 13.9 | 88 | 14.2 | 19 | 14.7 |
|  | No | 566 | 86.8 | 241 | 87.0 | 46 | 86.8 | 1215 | 86.1 | 533 | 85.8 | 110 | 85.3 |
| **Exposure to COVID-19** |  |  |  |  |  |  |  |  |  |  |  |  |  |
| Perceived exposure to COVID-19 | Low | 205 | 31.8 | 72 | 25.9 | 11 | 20.8 | 442 | 31.7 | 164 | 26.2 | 26 | 20.2 |
|  | High | 439 | 68.2 | 206 | 74.1 | 42 | 79.2 | 953 | 68.3 | 461 | 73.8 | 103 | 79.8 |
| Has been tested positive for COVID-19 | No | 491 | 76.1 | 210 | 75.5 | 41 | 77.4 | 1056 | 75.5 | 467 | 74.7 | 95 | 73.6 |
|  | Yes | 154 | 23.9 | 68 | 24.5 | 12 | 22.6 | 342 | 24.5 | 158 | 25.3 | 34 | 26.4 |
| Any relatives hospitalised for COVID-19 | No | 563 | 87.3 | 241 | 86.7 | 42 | 79.2 | 1224 | 87.6 | 543 | 86.9 | 103 | 79.8 |
|  | Yes | 82 | 12.7 | 37 | 13.3 | 11 | 20.8 | 174 | 12.4 | 82 | 13.1 | 26 | 20.2 |
| Complete vaccination COVID-19 | Yes | 468 | 73.1 | 207 | 75.3 | 37 | 69.8 | 977 | 70.4 | 442 | 71.5 | 89 | 69 |
|  | No | 172 | 26.9 | 68 | 24.7 | 16 | 30.2 | 411 | 29.6 | 176 | 28.5 | 40 | 31 |
| **Health comorbidities** |  |  |  |  |  |  |  |  |  |  |  |  |  |
| Post-traumatic stress disorder | No<sup>2</sup> | 555 | 91.3 | 225 | 86.2 |  |  | 1186 | 90.2 | 487 | 84 |  |  |
|  | Yes<sup>3</sup> | 53 | 8.7 | 36 | 13.8 |  |  | 129 | 9.8 | 93 | 16 |  |  |
| Depressive symptoms | Any to mild<sup>4</sup> | 486 | 78.9 | 202 | 76.5 | 10 | 18.9 | 1032 | 77.5 | 430 | 73.3 | 23 | 17.8 |
|  | Moderate to severe<sup>5</sup> | 130 | 21.1 | 62 | 23.5 | 43 | 81.1 | 299 | 22.5 | 157 | 26.7 | 106 | 82.2 |
| Anxiety symptoms | No<sup>6</sup> | 283 | 46.1 | 112 | 41.9 | 1 | 1.9 | 595 | 44.9 | 235 | 39.4 | 2 | 1.6 |
|  | Yes<sup>7</sup> | 331 | 53.9 | 155 | 58.1 | 52 | 98.1 | 729 | 55.1 | 362 | 60.6 | 127 | 98.4 |
| Burnout | No | 491 | 81.8 | 201 | 78.2 | 23 | 46.9 | 1064 | 82.6 | 442 | 78.4 | 56 | 47.5 |
|  | Yes<sup>8</sup> | 109 | 18.2 | 56 | 21.8 | 26 | 53.1 | 224 | 17.4 | 122 | 21.6 | 62 | 52.5 |
| Resilience | Yes<sup>9</sup> | 437 | 71.4 | 191 | 72.9 | 25 | 49.0 | 925 | 70.1 | 412 | 70.9 | 63 | 51.2 |
|  | No<sup>10</sup> | 175 | 28.6 | 71 | 27.1 | 26 | 51.1 | 395 | 29.9 | 169 | 29.1 | 60 | 48.8 |
| Experiencing a traumatic event<sup>11</sup> | No | 591 | 91.9 | 252 | 90.6 | 45 | 84.9 | 1289 | 92.3 | 569 | 91 | 109 | 84.5 |
|  | Yes | 52 | 8.1 | 26 | 9.4 | 8 | 15.1 | 107 | 7.7 | 56 | 9 | 20 | 15.5 |
| History of psychotropic treatment<sup>12</sup> | No | 424 | 69.7 | 167 | 63.7 | 20 | 40.8 | 896 | 67.8 | 360 | 61.2 | 46 | 38.3 |
|  | Yes | 184 | 30.3 | 95 | 36.3 | 29 | 59.2 | 425 | 32.2 | 228 | 38.8 | 74 | 61.7 |
| History of major lifetime somatic health problems | No | 384 | 60.7 | 145 | 53.9 | 22 | 44.0 | 846 | 61.6 | 318 | 52.6 | 50 | 41.3 |
|  | Yes | 249 | 39.3 | 124 | 46.1 | 28 | 56.0 | 528 | 38.4 | 286 | 47.4 | 71 | 58.7 |
| Psychological support related to the pandemic | No | 557 | 87.4 | 237 | 86.5 | 46 | 88.5 | 1202 | 87.02 | 533 | 86.5 | 111 | 88.8 |

|  | Raw data |  |  |  |  |  | Weighted data |  |  |  |  |  |
| --- | --- | --- | --- | --- | --- | --- | --- | --- | --- | --- | --- | --- |
|  | Total |  | Occupational stigma |  | PTSD symptoms |  | Total |  | Occupational stigma |  | PTSD symptoms |  |
|  | n = 655 |  | n = 278 |  | n = 53 |  | n = 1418 |  | n = 625 |  | n = 129 |  |
|  | n | % | n | % | n | % | n | % | n | % | n | % |
| Yes | 80 | 12.6 | 37 | 13.5 | 6 | 11.5 | 177 | 12.8 | 83 | 13.5 | 14 | 11.2 |

<sup>1</sup>Since the beginning of the pandemic
<sup>2</sup>PCL-5 score <33; <sup>3</sup>PCL-5 score ≥33
<sup>4</sup>PHQ-9 score <10; <sup>5</sup>PHQ-9 score ≥10
<sup>6</sup>GAD-7 score <10; <sup>7</sup>GAD-7 score ≥10
<sup>8</sup>MBI score: EE>29 and PA≤33 or EE>29 and DP>11; EE: emotional exhaustion; PA: personal accomplishment; DP: depersonalization
<sup>9</sup>CD-RISC-10 score ≥22; <sup>10</sup>CD-RISC-10 score <22
<sup>11</sup>in the 12 last months
<sup>12</sup>for more than 6 months
n: Number of individuals; %: Frequencies for each modality; PHQ-9: 9 items Health-Patient Questionnaire; PCL-5: Posttraumatic Stress Disorder Checklist; GAD-7: Generalized Anxiety Disorder Scale; MBI: Maslach Burnout Inventory test; CD-RISC-10: Connor-Davidson Resilience Scale-10

Univariate associations between covariates and PTSD symptoms and stigma, separately, on weighted and imputed data, are presented in Table 2. Covariates associated with PTSD symptoms were included to calculate propensity scores, as well as covariates associated with neither PTSD symptoms nor stigma. The univariate association between occupational stigma and PTSD symptoms was estimated by a crude OR = 4.05, 95% CI [2.03;8.07].

**Table 2.**
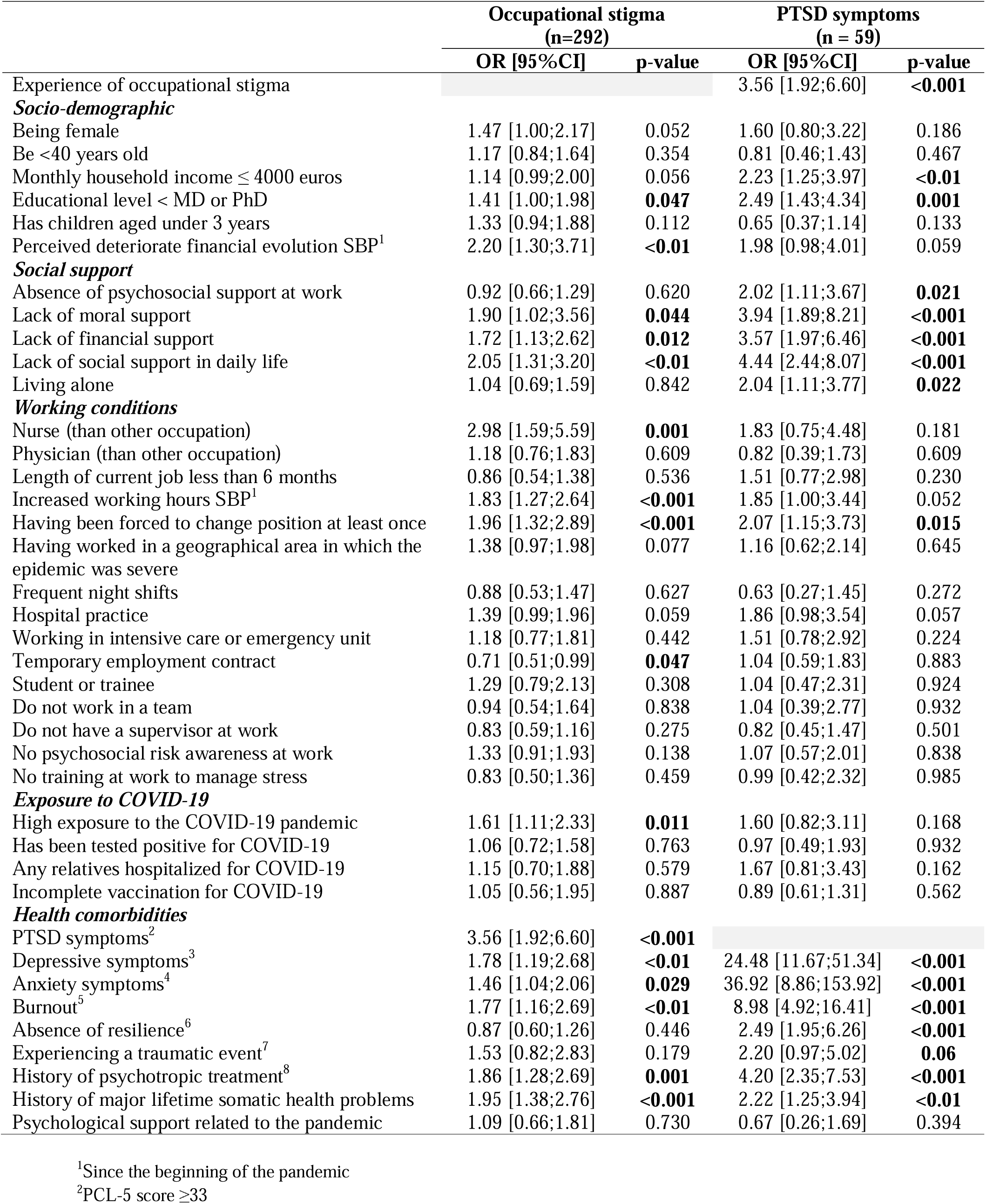
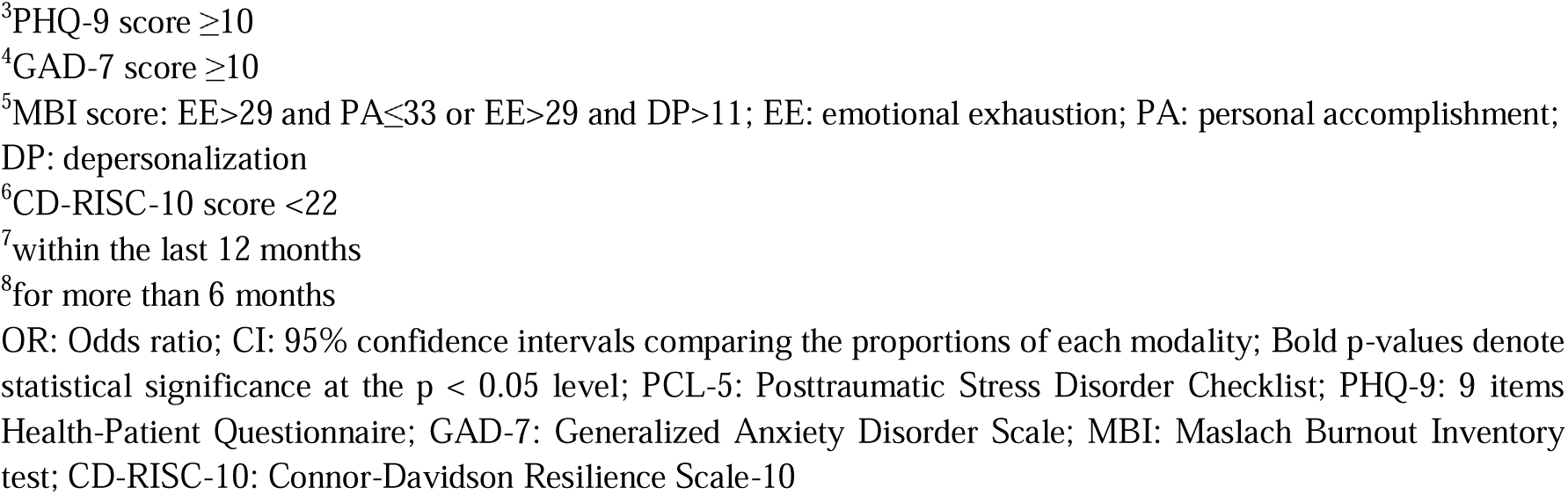
Univariate associations between 1) experience of perceived occupational stigma and PTSD symptoms, 2) covariates and stigma and PTSD symptoms, separately, on weighted and imputed data

|  | Occupational stigma<br>(n=292) |  | PTSD symptoms<br>(n = 59) |  |
| --- | --- | --- | --- | --- |
|  | OR [95%CI] | p-value | OR [95%CI] | p-value |
| Experience of occupational stigma |  |  | 3.56 [1.92;6.60] | <0.001 |
| ***Socio-demographic*** |  |  |  |  |
| Being female | 1.47 [1.00;2.17] | 0.052 | 1.60 [0.80;3.22] | 0.186 |
| Be <40 years old | 1.17 [0.84;1.64] | 0.354 | 0.81 [0.46;1.43] | 0.467 |
| Monthly household income ≤ 4000 euros | 1.14 [0.99;2.00] | 0.056 | 2.23 [1.25;3.97] | <0.01 |
| Educational level < MD or PhD | 1.41 [1.00;1.98] | **0.047** | 2.49 [1.43;4.34] | **0.001** |
| Has children aged under 3 years | 1.33 [0.94;1.88] | 0.112 | 0.65 [0.37;1.14] | 0.133 |
| Perceived deteriorate financial evolution SBP<sup>1</sup> | 2.20 [1.30;3.71] | <0.01 | 1.98 [0.98;4.01] | 0.059 |
| ***Social support*** |  |  |  |  |
| Absence of psychosocial support at work | 0.92 [0.66;1.29] | 0.620 | 2.02 [1.11;3.67] | **0.021** |
| Lack of moral support | 1.90 [1.02;3.56] | **0.044** | 3.94 [1.89;8.21] | <0.001 |
| Lack of financial support | 1.72 [1.13;2.62] | **0.012** | 3.57 [1.97;6.46] | <0.001 |
| Lack of social support in daily life | 2.05 [1.31;3.20] | <0.01 | 4.44 [2.44;8.07] | <0.001 |
| Living alone | 1.04 [0.69;1.59] | 0.842 | 2.04 [1.11;3.77] | **0.022** |
| ***Working conditions*** |  |  |  |  |
| Nurse (than other occupation) | 2.98 [1.59;5.59] | **0.001** | 1.83 [0.75;4.48] | 0.181 |
| Physician (than other occupation) | 1.18 [0.76;1.83] | 0.609 | 0.82 [0.39;1.73] | 0.609 |
| Length of current job less than 6 months | 0.86 [0.54;1.38] | 0.536 | 1.51 [0.77;2.98] | 0.230 |
| Increased working hours SBP<sup>1</sup> | 1.83 [1.27;2.64] | <0.001 | 1.85 [1.00;3.44] | 0.052 |
| Having been forced to change position at least once | 1.96 [1.32;2.89] | <0.001 | 2.07 [1.15;3.73] | **0.015** |
| Having worked in a geographical area in which the epidemic was severe | 1.38 [0.97;1.98] | 0.077 | 1.16 [0.62;2.14] | 0.645 |
| Frequent night shifts | 0.88 [0.53;1.47] | 0.627 | 0.63 [0.27;1.45] | 0.272 |
| Hospital practice | 1.39 [0.99;1.96] | 0.059 | 1.86 [0.98;3.54] | 0.057 |
| Working in intensive care or emergency unit | 1.18 [0.77;1.81] | 0.442 | 1.51 [0.78;2.92] | 0.224 |
| Temporary employment contract | 0.71 [0.51;0.99] | **0.047** | 1.04 [0.59;1.83] | 0.883 |
| Student or trainee | 1.29 [0.79;2.13] | 0.308 | 1.04 [0.47;2.31] | 0.924 |
| Do not work in a team | 0.94 [0.54;1.64] | 0.838 | 1.04 [0.39;2.77] | 0.932 |
| Do not have a supervisor at work | 0.83 [0.59;1.16] | 0.275 | 0.82 [0.45;1.47] | 0.501 |
| No psychosocial risk awareness at work | 1.33 [0.91;1.93] | 0.138 | 1.07 [0.57;2.01] | 0.838 |
| No training at work to manage stress | 0.83 [0.50;1.36] | 0.459 | 0.99 [0.42;2.32] | 0.985 |
| ***Exposure to COVID-19*** |  |  |  |  |
| High exposure to the COVID-19 pandemic | 1.61 [1.11;2.33] | **0.011** | 1.60 [0.82;3.11] | 0.168 |
| Has been tested positive for COVID-19 | 1.06 [0.72;1.58] | 0.763 | 0.97 [0.49;1.93] | 0.932 |
| Any relatives hospitalized for COVID-19 | 1.15 [0.70;1.88] | 0.579 | 1.67 [0.81;3.43] | 0.162 |
| Incomplete vaccination for COVID-19 | 1.05 [0.56;1.95] | 0.887 | 0.89 [0.61;1.31] | 0.562 |
| ***Health comorbidities*** |  |  |  |  |
| PTSD symptoms<sup>2</sup> | 3.56 [1.92;6.60] | <0.001 |  |  |
| Depressive symptoms<sup>3</sup> | 1.78 [1.19;2.68] | <0.01 | 24.48 [11.67;51.34] | <0.001 |
| Anxiety symptoms<sup>4</sup> | 1.46 [1.04;2.06] | **0.029** | 36.92 [8.86;153.92] | <0.001 |
| Burnout<sup>5</sup> | 1.77 [1.16;2.69] | <0.01 | 8.98 [4.92;16.41] | <0.001 |
| Absence of resilience<sup>6</sup> | 0.87 [0.60;1.26] | 0.446 | 2.49 [1.95;6.26] | <0.001 |
| Experiencing a traumatic event<sup>7</sup> | 1.53 [0.82;2.83] | 0.179 | 2.20 [0.97;5.02] | **0.06** |
| History of psychotropic treatment<sup>8</sup> | 1.86 [1.28;2.69] | **0.001** | 4.20 [2.35;7.53] | <0.001 |
| History of major lifetime somatic health problems | 1.95 [1.38;2.76] | <0.001 | 2.22 [1.25;3.94] | <0.01 |
| Psychological support related to the pandemic | 1.09 [0.66;1.81] | 0.730 | 0.67 [0.26;1.69] | 0.394 |

<sup>1</sup>Since the beginning of the pandemic
<sup>2</sup>PCL-5 score ≥33
<sup>3</sup>PHQ-9 score $\geq 10$
<sup>4</sup>GAD-7 score $\geq 10$
<sup>5</sup>MBI score: $EE > 29$ and $PA \leq 33$ or $EE > 29$ and $DP > 11$ ; EE: emotional exhaustion; PA: personal accomplishment; DP: depersonalization
<sup>6</sup>CD-RISC-10 score $< 22$
<sup>7</sup>within the last 12 months
<sup>8</sup>for more than 6 months
OR: Odds ratio; CI: 95% confidence intervals comparing the proportions of each modality; Bold p-values denote statistical significance at the $p < 0.05$ level; PCL-5: Posttraumatic Stress Disorder Checklist; PHQ-9: 9 items Health-Patient Questionnaire; GAD-7: Generalized Anxiety Disorder Scale; MBI: Maslach Burnout Inventory test; CD-RISC-10: Connor-Davidson Resilience Scale-10

After creating propensity scores (PS), covariate balance across groups of occupational stigma was tested using ASMD and KS. Propensity-score weighted descriptive statistics by occupational stigma are presented in Supplementary Table 1.

Inverse probability weighting (IPW) was adjusted on: gender, age, monthly household income, educational level, having children, psychosocial support at work, moral support, financial support, social support in daily life, length of current job, having been forced to change position, having worked in a geographical area in which the epidemic was severe, night shifts, place of practice, working in intensive care or emergency unit, student or trainee, work in a team, supervisor at work, psychosocial risk awareness at work, training at work to manage stress, has been tested positive for COVID-19, relatives hospitalized for COVID-19, vaccination for COVID-19, depressive symptoms, anxiety symptoms, burnout, resilience, experiencing a traumatic event, history of psychotropic treatment, history of major lifetime somatic health problems and psychological support related to the pandemic.

Covariates balance before and after adjustment is presented in Supplementary Fig1. It showed a good fit. Only the variables “living alone” and “work in a team” were removed as they were not well balanced.

After IPW adjustment, experience of perceived occupational stigma was significantly associated with PTSD symptoms with an adjusted ORa=2.16; 95% CI [1.09;4.25].

The sensitivity analysis showed that 189 of the 532 HCWs (35.5%) reported experience of occupational stigma when changing its definition. Univariate associations between sigma and PSTD, and between cofactors and PTSD and stigma separately, are presented in Supplementary table 2. Propensity-score weighted descriptive statistics by experience of perceived occupational stigma are presented in Supplementary Table 3. Covariate balance before and after adjustment is presented in Supplementary Fig2., showing a good fit. In this sensibility analysis, 11 variables (psychosocial support at work, living alone, having worked in a geographical area in which the epidemic was severe, work in a team, psychosocial risk awareness at work, has been tested positive for COVID-19, incomplete vaccination for COVID-19, depressive symptoms, burnout, resilience and history of psychotropic treatment for more than 6 months) were deleted because they didn’t balance well. Compared with the main analysis, the sensitivity analysis led us to remove more variables (11 vs. 2) in the covariate balance. Reducing the sample size when recoding stigma in the sensitivity analysis (from 655 to 532 individuals) may have created greater homogeneity and therefore less variability for some variables.

After IPW adjustment, this sensitivity analysis estimated an association between stigma and PTSD very similar to that of our main analysis, with an adjusted ORa = 2.17; 95% CI [1.04, 4.52].

## Discussion

One year after the onset of the COVID-19 pandemic in France, 8.7% of our study population exhibited symptoms of PTSD, while 44.8% reported experience of occupational stigmatization.

We identified a prevalence of PTSD much lower than the 26% reported in a recently published meta-analysis of 161 studies including 341,014 HCWs worldwide (Huang et al., 2024). Its authors highlighted a high between-study heterogeneity, with individual study estimates ranging from 2% to 67%. Higher PTSD prevalence found in other studies may be due to the selective inclusion of healthcare workers who were more exposed (working in emergency and intensive care units) (Bahadirli & Sagaltici, 2021; Carola et al., 2022). It may also be due to the professions studied. Indeed, some studies included mostly nurses and nursing assistants, who are more disadvantaged than doctors (Esteban-Sepúlveda et al., 2022) in terms of educational level, financial status (Bonzini et al., 2022; Costa et al., 2023) and who may have less access to mental health care. In addition, nurses and nursing assistants may have been more on the emotional front line with patients and their families and therefore more exposed to potentially traumatic events (patient suffering or death, family worries and grief).

Nearly half of those surveyed said they had been stigmatized because of their profession one year into the pandemic. Although cited in numerous WHO recommendations on the mental health of HCWs during the COVID-19 pandemic (Abdul-Rahim et al., 2022), the frequency of their experiences of stigmatization and rejection has rarely been estimated. It seems particularly high in a country like France where the population has massively adhered to COVID-19-related policies. When reported in the media, these situations of stigmatization, distancing, or rejection have also aroused strong disapproval in France. However, they reflected the degree of fear among the population in the face of a dangerous new virus. If new pandemics of transmissible agents were to occur, there is nothing to say that such reactions of fear and rejection of caregivers would be rarer; quite the contrary, since fake news and the manipulation of public opinion in alternative media and on social networks have since multiplied.

We found that such a stigma was significantly associated with an increased risk of PTSD symptoms, after controlling numerous sociodemographic characteristics, social support, working conditions, COVID-19 exposure, and comorbidities through propensity score estimation. Such an association has been observed in others studies adjusted on sociodemographic characteristics and working conditions (Narita et al., 2022; Van Wert et al., 2022) or mental health and exposure to COVID-19 (Zhou et al., 2022), but –to our knowledge – never for all of them.

Our study is unique in demonstrating that experiencing occupational stigmatization itself is associated with PTSD symptoms one year after the onset of the COVID-19 pandemic, even after accounting for social characteristics and comorbidities. The results of our sensitivity analysis highlighted also a relationship between stigma and PTSD even after accounting for social characteristics.

Our study has some limitations. First, missing data needs to be considered. We attempted to mitigate this problem by imputing missing data to reduce simulation error. However, given the unique distribution of our sample, which consists primarily of physicians with higher incomes compared to nurses or nursing assistants, it is difficult to generalize our results to all healthcare providers in France. Second, the voluntary recruitment process may have introduced selection bias. Individuals with less exposure to COVID-19 pandemic and who were coping well may have felt they did not meet the participation criteria because the survey focused on the mental health of HCWs. Conversely, those with higher exposure and poorer mental health may have been more likely to participate (Grievink et al., 2006). Third, since our survey is cross-sectional (notably, the duration of PTSD symptoms remains unknown, as does their chronology with stigmatization experiences), the interpretation of the results must be cautious and does not allow for a causal link to be concluded. Indeed, it cannot be formally excluded that among the 8.7% of HCWs with symptoms of PTSD, it is these symptoms which could increase their susceptibility to feelings of stigmatization. Fourth, the experience of occupational stigma was measured using a single question, which limits the comparability and generalizability of the results compared to using a more detailed scale. In the context of the COVID-19 pandemic, two studies, conducted in developing countries, used stigma measurement scales adapted from those previously developed for other pandemics. Both found higher prevalences of stigma among HCWs than in our study: Giri et al. adapted the Middle East respiratory syndrome coronavirus (MERS-CoV) stigma scale and found a stigma prevalence of 35% in Népal (Giri et al., 2022); and Cénat et al. adapted The Stigmatization related to Ebola virus disease (EVD) scale to COVID-19 and found a stigma prevalence of 47% in Eastern Congo (Cénat et al., 2022). However, the study conducted by Asnakew et al. in Ethiopia, which, like ours, used a single question on perceived stigma, found a lower prevalence than our study (24%), but an association between stigma and PTSD close to our results (a0R=1.97) (Asnakew et al., 2021). The use of a single question may underestimate the prevalence of stigma (including in our study), but comparisons remain difficult between such different contexts.

Our study had several strengths. First, to our knowledge, this is a pioneering study examining the association between occupational stigma experienced by HCWs and their PTSD symptoms, adjusting for the large number of possible cofounders or associated factors available in the PsyCOVer survey: sociodemographic characteristics, socioeconomic status, social support, working conditions, exposure to COVID-19, and health comorbidities, The results of the sensitivity analysis strengthened our conclusions as they were robust to changes in stigmatization categories. Second, we used data calibration with the margin adjustment method and included survey weights in the logistic regression model, which allowed us to reduce selection bias. Importantly, the use of a propensity scores instead of a classical regression model allowed for model specification verification, separating study design from study analysis, avoiding the temptation to keep tweaking the model until the desired association is obtained, enabling the inclusion of an unlimited number of relevant covariates in the model, and facilitating the explicit comparison of covariate distributions between the two groups (Austin, 2011).

## Conclusion

Our findings suggest that healthcare workers who have experienced occupational stigma during the first year of the COVID-19 pandemic may be at increased risk for PTSD symptoms, independent of many characteristics known to influence both occupational stigma and PTSD. For future research on the mental health of HCWs, our study highlights the importance of examining in more detail the contexts and circumstances of their experiences of occupational stigmatization, investigating the direction of the association between them and PTSD, and focusing more on potential mediating factors of this relationship.

Preventing the onset of PTSD symptoms among healthcare workers following epidemics such as COVID-19 is of paramount importance. Occupational stigma may be a potential occupational risk factor that can be targeted to improve the working conditions and mental health of healthcare workers. These findings highlight the importance of protecting healthcare workers from occupational stigma through support and counseling interventions for exposed professionals during pandemics and periods of high pressure on healthcare facilities. Furthermore, implementing information and prevention campaigns among the general population against the stigma of healthcare workers is crucial. These campaigns should involve public education and the active participation of government authorities.

## Data Availability

The data that support the findings of this study are available from the National Institute of Health and Medical Research (Inserm, "Institut national de la sante et de la recherche medicale") but restrictions apply to the availability of these data, which were used under license for the current study, and so are not publicly available. Data are, however, available from the authors upon reasonable request and with permission of the Inserm.

## Acknowledgements

The authors are most grateful to all the study participants for their involvement, especially given the very difficult context for them.

## Author contributions: CRediT

Charline VINCENT: Conceptualization, Methodology, Software, Formal analysis, Writing - Original Draft; Roberto MEDIAVILLA, Honor SCARLETT, Wissam EL-HAGE and Pierre CHAUVIN: Review & Editing; Cécile VUILLERMOZ: Data Curation, Review & Editing, Supervision, Project administration, Funding acquisition.

## Funding sources

The data collection of the PsyCOVer project was funded by the Agence Nationale de la Recherche (ANR) grant proposal ‘RA-Covid-19’. This study is part of the doctoral work of the first author Charline VINCENT, which is funded by a doctoral contract from Sorbonne University. The work of Roberto Mediavilla was supported by the Instituto de Salud Carlos III and the European Regional Development Fund (CD22/00061).

## Conflict of Interest statement

The authors declare no conflict of interest.

## Ethics statements

The authors assert that all procedures contributing to this work comply with the ethical standards of the relevant national and institutional committees on human experimentation and with the Helsinki Declaration of 1975, as revised in 2008. The study protocol was approved by the Sorbonne University Ethical Committee (No 2020-CER-2020-27) and was reported to the French Commission on Information Technology and Liberties, CNIL (N°2222413, 20-05-2021). All methods were performed in accordance with the relevant guidelines and regulations. Participants could contact the research team to discuss the findings of the study which were in the form of aggregated results as data will be anonymous and non-identifiable to the researcher. Written informed consent was obtained from all participants.

## Data Availability statement

The data that support the findings of this study are available from the National Institute of Health and Medical Research (Inserm, “Institut national de la santé et de la recherche médicale”) but restrictions apply to the availability of these data, which were used under license for the current study, and so are not publicly available. Data are, however, available from the authors upon reasonable request and with permission of the Inserm.

## Appendices

Table 1. Description of the study population

Table 2. Univariate associations between 1) experience of perceived occupational stigma and PTSD symptoms, 2) covariates and stigma and PTSD symptoms, separately, on weighted and imputed data

## Notes

### Competing Interest Statement

The authors have declared no competing interest.

### Author Declarations

The authors assert that all procedures contributing to this work comply with the ethical standards of the relevant national and institutional committees on human experimentation and with the Helsinki Declaration of 1975, as revised in 2008. The study protocol was approved by the Sorbonne University Ethical Committee (No 2020-CER-2020-27) and was reported to the French Commission on Information Technology and Liberties, CNIL (Number 2222413, 20-05-2021). All methods were performed in accordance with the relevant guidelines and regulations. Participants could contact the research team to discuss the findings of the study which were in the form of aggregated results as data will be anonymous and non-identifiable to the researcher. Written informed consent was obtained from all participants.

