## Supplementary material for "Occupational stigma and post-traumatic stress disorder among healthcare workers"

**Supplementary Table 1.** Propensity-score weighted descriptive statistics for experience of perceived occupational stigma, on weighted and imputed data.

|  | Mean ASMD | Max ASMD | Mean  KS-test statistic | Max  KS-test statistic |
| --- | --- | --- | --- | --- |
| Being female | 0.0578 | 0.0743 | 0.0236 | 0.0303 |
| Be <40 years old | 0.0589 | 0.0527 | 0.0294 | 0.0263 |
| Monthly household income ≤ 4000 euros | 0.0679 | 0.0735 | 0.0334 | 0.0360 |
| Educational level < MD or PhD | 0.0453 | 0.0595 | 0.0226 | 0.0296 |
| Has children aged under 3 years | 0.0578 | 0.0857 | 0.0279 | 0.0414 |
| Absence of psychosocial support at work | -0.0114 | 0.0430 | 0.0115 | 0.0214 |
| Lack of moral support | 0.0412 | 0.0791 | 0.0112 | 0.0215 |
| Lack of financial support | 0.1477 | 0.1245 | 0.0617 | 0.0519 |
| Lack of social support in daily life | 0.1242 | 0.1178 | 0.0485 | 0.0462 |
| Living alone | 0.0533 | 0.0357 | 0.0218 | 0.0146 |
| Length of current job less than 6 months | -0.0207 | 0.0328 | 0.0086 | 0.0114 |
| Having been forced to change position at least once | 0.1469 | 0.0949 | 0.0640 | 0.0413 |
| Having worked in a geographical area in which the epidemic was severe | 0.0266 | 0.0979 | 0.0152 | 0.0457 |
| Frequent night shifts | -0.0318 | 0.0487 | 0.0109 | 0.0166 |
| Hospital practice | 0.1049 | 0.0553 | 0.0492 | 0.0259 |
| Working in intensive care or emergency unit | 0.0287 | 0.0640 | 0.0138 | 0.0246 |
| Student or trainee | 0.0488 | 0.0462 | 0.0152 | 0.0144 |
| Do not work in a team | -0.0415 | 0.0590 | 0.0118 | 0.0168 |
| Do not have a supervisor at work | -0.0146 | 0.0596 | 0.0126 | 0.0289 |
| No psychosocial risk awareness at work | 0.0285 | 0.1141 | 0.0190 | 0.0519 |
| No training at work to manage stress | -0.0656 | 0.0936 | 0.0228 | 0.0324 |
| Has been tested positive for COVID-19 | 0.0196 | 0.0600 | 0.0108 | 0.0259 |
| Any relatives hospitalized for COVID-19 | 0.0366 | 0.0508 | 0.0127 | 0.0170 |
| Incomplete vaccination for COVID-19 | 0.0085 | 0.0743 | 0.0104 | 0.0338 |
| Depressive symptoms^1^ | 0.1286 | 0.0838 | 0.0542 | 0.0352 |
| Anxiety symptoms^2^ | 0.0929 | 0.1052 | 0.0459 | 0.0520 |
| Burnout^3^ | 0.1293 | 0.1144 | 0.0503 | 0.0450 |
| Absence of resilience^4^ | -0.0262 | 0.0765 | 0.0172 | 0.0350 |
| Experiencing a traumatic event^5^ | 0.0089 | 0.0761 | 0.0069 | 0.0210 |
| History of psychotropic treatment^6^ | 0.1141 | 0.1383 | 0.0529 | 0.0644 |
| History of major lifetime somatic health problems | 0.1851 | 0.1571 | 0.0889 | 0.0753 |
| Psychological support related to the pandemic | 0.0043 | 0.0403 | 0.0082 | 0.0137 |

^1^PHQ-9 score ≥10

^2^GAD-7 score ≥10

^3^MBI score: EE>29 and PA≤33 or EE>29 and DP>11; EE: emotional exhaustion; PA: personal accomplishment; DP: depersonalization

^4^CD-RISC-10 score <22

^5^in the 12 last months

^6^for more than 6 months

ASMD: Absolute Standard Mean Difference; KS-test: Kolmogorov-Smirnov test statistic; PHQ-9: 9 items Health-Patient Questionnaire; GAD-7: Generalized Anxiety Disorder Scale; MBI: Maslach Burnout Inventory test; CD-RISC-10: Connor-Davidson Resilience Scale-10

**Supplementary Fig1.** Covariate balance before and after adjustment*.*

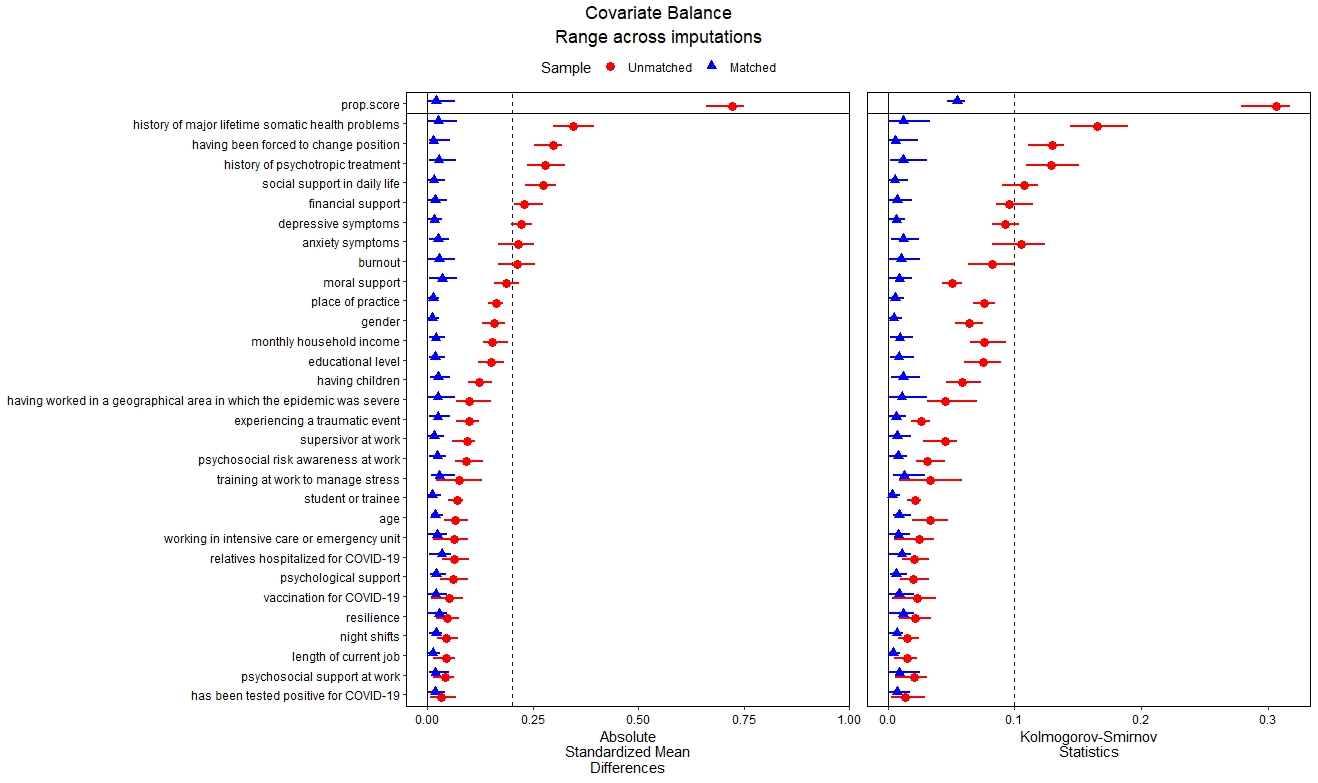

**Supplementary table 2.** Sensitivity analysis of univariate associations between 1) experience of perceived occupational stigma and PTSD symptoms, 2) covariates and stigma and PTSD symptoms, separately, on weighted and imputed data

|  | **Occupational stigma**  **(n=189)** | | | | **PTSD symptoms**  **(n = 42)** | |
| --- | --- | --- | --- | --- | --- | --- |
|  | **OR [_95%_CI]** | **p-value** | | **OR [_95%_CI]** | | **p-value** |
| Experience of occupational stigma |  | |  |  | |  |
| ***Socio-demographic*** |  | |  |  | |  |
| Being female | 1.91 [1.18;3.10] | | **<0.01** | 1.34 [0.61;2.97] | | 0.465 |
| Be <40 years old | 0.86 [0.58;1.26] | | 0.432 | 1.09 [0.57;2.11] | | 0.792 |
| Monthly household income ≤ 4000 euros | 1.30 [0.87;1.94] | | 0.200 | 1.75 [0.91;3.40] | | 0.096 |
| Educational level < MD or PhD | 1.44 [0.98;2.12] | | 0.063 | 1.76 [0.93;3.34] | | 0.081 |
| Has children aged under 3 years | 1.50 [1.00;2.25] | | **0.050** | 0.87 [0.45;1.68] | | 0.672 |
| Perceived deteriorate financial evolution SBP^1^ | 2.41 [1.34;4.33] | | **<0.01** | 1.79 [0.76;4.22] | | 0.183 |
| ***Social support*** |  | |  |  | |  |
| Absence of psychosocial support at work | 0.88 [0.59;1.30] | | 0.512 | 1.44 [0.74;2.80] | | 0.284 |
| Lack of moral support | 1.77 [0.86;3.65] | | 0.120 | 5.54 [2.39;12.98] | | **<0.001** |
| Lack of financial support | 1.85 [1.15;2.98] | | **0.011** | 2.32 [1.16;4.64] | | **0.018** |
| Lack of social support in daily life | 1.93 [1.18;3.16] | | **<0.01** | 4.43 [2.23;8.82] | | **<0.001** |
| Living alone | 0.98 [0.60;1.59] | | 0.934 | 1.15 [0.54;2.46] | | 0.722 |
| ***Working conditions*** |  | |  |  | |  |
| Nurse (than other occupation) | 3.17 [1.55;6.43] | | **<0.001** | 2.47 [0.80;7.67] | | 0.117 |
| Physician (than other occupation) | 1.21 [0.71;2.05] | | 0.485 | 1.29 [0.50;3.35] | | 0.601 |
| Length of current job less than 6 months | 0.73 [0.41;1.29] | | 0.279 | 1.26 [0.57;2.79] | | 0.569 |
| Increased working hours SBP^1^ | 2.25 [1.44;3.51] | | **<0.001** | 2.67 [1.25;5.69] | | **0.011** |
| Having been forced to change position at least once | 2.42 [1.56;3.76] | | **<0.001** | 2.84 [1.45;5.56] | | **<0.01** |
| Having worked in a geographical area in which the epidemic was severe | 1.14 [0.75;1.74] | | 0.545 | 0.91 [0.45;1.83] | | 0.787 |
| Frequent night shifts | 0.82 [0.46;1.46] | | 0.507 | 0.92 [0.39;2.20] | | 0.857 |
| Hospital practice | 1.36 [0.91;2.02] | | 0.134 | 1.35 [0.67;2.73] | | 0.399 |
| Working in intensive care or unit | 1.13 [0.69;1.85] | | 0.615 | 1.95 [0.92;4.12] | | 0.080 |
| Temporary employment contract | 0.69 [0.47;1.02] | | 0.061 | 1.00 [0.52;1.92] | | 1.000 |
| Student or trainee | 1.53 [0.84;2.78] | | 0.166 | 1.21 [0.45;3.25] | | 0.700 |
| Do not work in a team | 0.96 [0.50;1.83] | | 0.899 | 1.32 [0.45;3.39] | | 0.611 |
| Do not have a supervisor at work | 0.84 [0.57;1.24] | | 0383 | 0.81 [0.41;1.59] | | 0.531 |
| No psychosocial risk awareness at work | 1.01 [0.66;1.54] | | 0.974 | 1.51 [0.71;3.22] | | 0.289 |
| No training at work to manage stress | 0.75 [0.42;1.34] | | 0.325 | 0.91 [0.34;2.43] | | 0.843 |
| ***Exposure to COVID-19*** |  | |  |  | |  |
| High exposure to the COVID-19 pandemic | 2.18 [1.38;3.44] | | **<0.001** | 2.50 [1.04;5.98] | | **0.040** |
| Has been tested positive for COVID-19 | 1.07 [0.68;1.68] | | 0.763 | 0.72 [0.32;1.63] | | 0.426 |
| Any relatives hospitalized for COVID-19 | 1.27 [0.72;2.24] | | 0.410 | 2.29 [1.02;5.11] | | **0.044** |
| Incomplete vaccination for COVID-19 | 0.93 [0.60;1.44] | | 0.742 | 0.70 [0.33;1.45] | | 0.330 |
| ***Health comorbidities*** |  | |  |  | |  |
| PTSD symptoms^2^ | 4.05 [2.03;8.07] | | **<0.001** |  | |  |
| Depressive symptoms^3^ | 1.73 [1.08;2.75] | | **0.022** | 17.31 [7.95;37.68] | | **<0.001** |
| Anxiety symptoms^4^ | 1.77 [1.19;2.65] | | **<0.01** | 32.67 [7.2;148.3] | | **<0.001** |
| Burnout^5^ | 2.01 [1.24;3.24] | | **<0.01** | 6.61 [3.35;13.05] | | **<0.001** |
| Absence of resilience^6^ | 0.99 [0.64;1.52] | | 0.960 | 2.45 [1.26;4.76] | | **<0.01** |
| Experiencing a traumatic event^7^ | 1.36 [0.68;2.72] | | 0.380 | 2.28 [0.95;5.47] | | 0.065 |
| History of psychotropic treatment^8^ | 2.11 [1.38;3.22] | | **<0.001** | 3.03 [1.56;5.88] | | **<0.01** |
| History of major lifetime somatic health problems | 2.24 [1.51;3.34] | | **<0.001** | 1.85 [0.96;3.58] | | 0.067 |
| Psychological support related to the pandemic | 1.32 [0.75;2.34] | | 0.339 | 1.39 [0.56;3.46] | | 0.473 |

^1^Since the beginning of the pandemic

^2^PCL-5 score ≥33

^3^PHQ-9 score ≥10

^4^GAD-7 score ≥10

^5^MBI score: EE>29 and PA≤33 or EE>29 and DP>11; EE: emotional exhaustion; PA: personal accomplishment; DP: depersonalization

^6^CD-RISC-10 score <22

^7^in the 12 last months

^8^for more than 6 months

OR: Odds ratio; CI: 95% confidence intervals comparing the proportions of each modality; Bold p-values denote statistical significance at the p < 0.05 level; PCL-5: Posttraumatic Stress Disorder Checklist; PHQ-9: 9 items Health-Patient Questionnaire; GAD-7: Generalized Anxiety Disorder Scale; MBI: Maslach Burnout Inventory test; CD-RISC-10: Connor-Davidson Resilience Scale-10

**Supplementary Table 3.** Sensitivity analysis of propensity-score weighted descriptive statistics by experience of perceived occupational stigma, on weighted and imputed data.

|  | Mean ASMD | Max ASMD | Mean  KS-test statistic | | Max  KS-test statistic |
| --- | --- | --- | --- | --- | --- |
| Be <40 years old | -0.0704 | 0.0833 | 0.0351 | 0.0415 | |
| Monthly household income ≤ 4000 euros | 0.0329 | 0.0814 | 0.0168 | 0.0402 | |
| Educational level < MD or PhD | 0.0393 | 0.0393 | 0.0195 | 0.0195 | |
| Has children aged under 3 years | 0.0885 | 0.0948 | 0.0424 | 0.0454 | |
| Lack of moral support | 0.0157 | 0.0312 | 0.0047 | 0.0085 | |
| Lack of financial support | 0.1134 | 0.1435 | 0.0478 | 0.0606 | |
| Lack of social support in daily life | 0.0614 | 0.0766 | 0.0244 | 0.0303 | |
| Length of current job less than 6 months | 0.0166 | 0.0571 | 0.0069 | 0.0195 | |
| Increased working hours SBP^1^ | 0.1293 | 0.1293 | 0.0587 | 0.0587 | |
| Having been forced to change position at least once | 0.1657 | 0.1827 | 0.0735 | 0.0809 | |
| Frequent night shifts | -0.0554 | 0.0554 | 0.0189 | 0.0189 | |
| Hospital practice | 0.1044 | 0.1086 | 0.0493 | 0.0512 | |
| Working in intensive care or emergency unit | 0.0061 | 0.0061 | 0.0023 | 0.0023 | |
| Student or trainee | 0.0533 | 0.0533 | 0.0162 | 0.0162 | |
| Temporary employment contract | -0.0621 | 0.0882 | 0.0306 | 0.0434 | |
| Do not have a supervisor at work | -0.0494 | 0.0622 | 0.0239 | 0.0301 | |
| No training at work to manage stress | -0.0521 | 0.0965 | 0.0177 | 0.0331 | |
| High exposure to the COVID-19 pandemic | 0.1264 | 0.1314 | 0.0561 | 0.0584 | |
| Any relatives hospitalized for COVID-19 | 0.0281 | 0.0281 | 0.0093 | 0.0093 | |
| Anxiety symptoms^2^ | 0.1080 | 0.1531 | 0.0527 | 0.0748 | |
| Experiencing a traumatic event^3^ | -0.0386 | 0.0396 | 0.0102 | 0.0105 | |
| Psychological support related to the pandemic | -0.0030 | 0.0376 | 0.0074 | 0.0128 | |

^1^Since the beginning of the pandemic

^2^GAD-7 score ≥10

^3^in the 12 last months

GAD-7: Generalized Anxiety Disorder Scale; ASMD: Absolute Standard Mean Difference; KS: Kolmogorov-Smirnov test statistic

**Supplementary Fig2.** Covariate balance of sensitivity analysis before and after adjustment

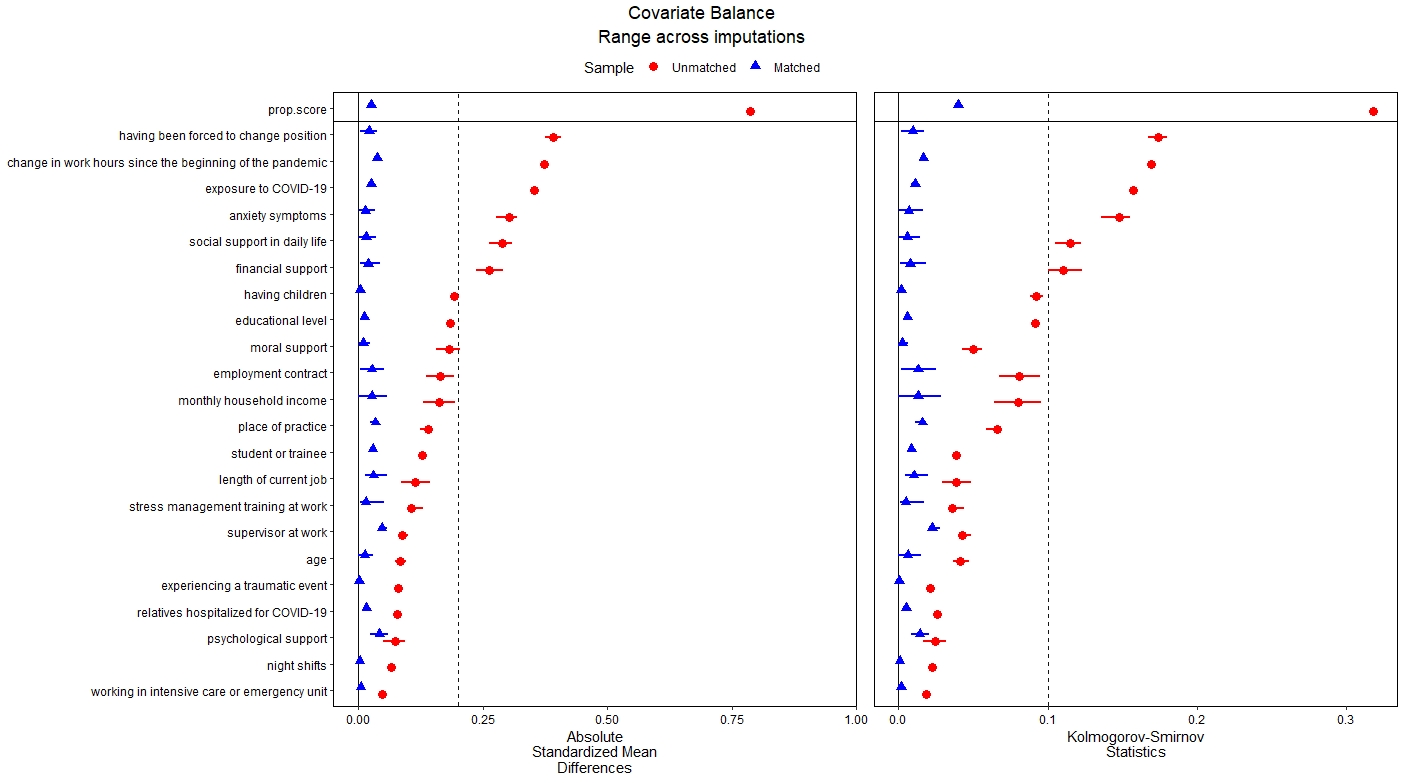
